# SARS-CoV-2 evolves increased infection elicited cell death and fusion in an immunosuppressed individual

**DOI:** 10.1101/2022.11.23.22282673

**Authors:** Gila Lustig, Yashica Ganga, Hylton Rodel, Houriiyah Tegally, Laurelle Jackson, Sandile Cele, Khadija Khan, Zesuliwe Jule, Kajal Reedoy, Farina Karim, Mallory Bernstein, Mahomed-Yunus S. Moosa, Derseree Archary, Tulio de Oliveira, Richard Lessells, Salim S. Abdool Karim, Alex Sigal

**Author notes:** Equal contribution (Gila Lustig, Yashica Ganga).

## Abstract

The milder clinical manifestations of Omicron infection relative to pre-Omicron SARS-CoV-2 raises the possibility that extensive evolution results in reduced pathogenicity. To test this hypothesis, we quantified induction of cell fusion and cell death in SARS-CoV-2 evolved from ancestral virus during long-term infection. Both cell fusion and death were reduced in Omicron BA.1 infection relative to ancestral virus. Evolved virus was isolated at different times during a 6-month infection in an immunosuppressed individual with advanced HIV disease. The virus isolated 16 days post-reported symptom onset induced fusogenicity and cell death at levels similar to BA.1. However, fusogenicity was increased in virus isolated at 6 months post-symptoms to levels intermediate between BA.1 and ancestral SARS-CoV-2. Similarly, infected cell death showed a graded increase from earlier to later isolates. These results may indicate that, at least by the cellular measures used here, evolution in long-term infection does not necessarily attenuate the virus.

## Introduction

The reduced incidence of severe disease reported with Omicron^1^ may result from the increasing immunity of the population because of vaccination and previous infections. Alternatively, the virus itself may have decreased its propensity to cause more severe disease^2-4^. If the ability of the Omicron virus itself to cause severe disease is attenuated independently of increased population immunity, the question arises of whether the virus is constrained to attenuate because of the evolutionary process through which variants emerge.

Mechanisms of variant formation may include reverse zoonosis^5-14^, the infection of an animal reservoir where the virus mutates to adapt to the new host species, then re-infection of a human host, or evolution in long-term infection in immunosuppressed individuals^15-27^. Evolution in long-term infection in immunosuppression is documented to occur in some people who are immunosuppressed because of advanced HIV disease^15-17,26^, defined as a CD4 T cell count < 200 cells/microliter in a person living with HIV.

Analysis of multiple long-term infections in immunosuppressed individuals has demonstrated recurrent mutations that are associated with escape from neutralizing antibodies^24^. However, mutations outside of spike are also common^28^ and may affect virus infection and cell-cell spread and may therefore impact pathogenicity^24^, with one possible outcome being that pathogenicity is reduced. In cell culture, viruses attenuate during long-term passaging, and such passaging is used to make live attenuated vaccines^29^.

SARS-CoV-2 infection can lead to disease in several ways^30^. One hallmark is the presence in the lung of syncytia, cells which underwent fusion through the interaction of the SARS-CoV-2 spike protein expressed on the infected cell surface with the human angiotensin converting enzyme 2 (ACE2) receptor on neighbouring cells^31-34^. Fusion results in multinucleated cells. This phenotype can be readily reproduced in cell culture infection with SARS-CoV-2^35-38^.

The sequence of events which leads to the ability of the virus to enter cells by binding ACE2 on the plasma membrane is also required for infected cells to efficiently fuse to other cells. SARS-CoV-2 spike has two subunits, S1 and S2. The S1 subunit binds the ACE2 receptor, while S2 mediates membrane fusion^39^. Spike contains an S1/S2 cleavage site predominantly cleaved by the cellular furin protease^32^. This cleavage allows further cleavage at the S2’ site mediated by the cellular serine protease TMPRSS2, activating the S2 subunit for fusion^39^. A cathepsin-dependent alternate pathway for viral entry exists and enables TMPRSS2-independent viral infection^32^. The Omicron BA.1 subvariant does not have efficient S1/S2 cleavage and predominantly uses the alternative pathway to infect^31,40,41^. Consequently, cell-cell fusion induced by BA.1 is lower than with ancestral SARS-CoV-2 and Delta variant infections^31^. Generally, pre-Omicron variants show enhanced fusogenicity compared to ancestral virus^42-45^, while Omicron subvariants BA.1 and BA.2 show decreased fusogenicity which correlates with decreased pathogenicity in hamster infections^31,44,46,47^. In the Omicron BA.4 and BA.5 subvariants, fusogenicity is higher relative to BA.1 and BA.2^44,46^, and this is associated with higher pathogenicity in hamsters^46^.

SARS-CoV-2 infection also leads to death of infected cells^48,49^ which can be recapitulated in cellular assays^50-52^. Cell death may initiate an inflammatory response and lead to the trafficking of immune cells to the site of inflammation (reviewed in^53,54^). This causes increased lung fluid, cellularity, and later scarring, and in turn leads to less oxygen being able to diffuse into the blood, resulting in respiratory distress (reviewed in^55^). A reduction in cell death upon infection would therefore be predicted to lead to lower levels of inflammation and therefore lower disease severity.

We have previously reported on the evolution of SARS-CoV-2 from ancestral virus infection in an individual who was immunosuppressed because of advanced HIV disease^15,16^. Here we used timelapse microscopy and flow cytometry assays to determine changes in cell fusion and cell death induced by SARS-CoV-2 as it evolves over half a year of continuous infection. As a reference, we show cell fusion and cell death with Omicron BA.1 infection and ancestral virus with the D614G substitution.

We observed that virus from relatively early in the infection induced cell fusion and death at similar levels to Omicron BA.1. However, the virus which evolved over 6 months of infection showed fusogenicity intermediate between BA.1 and D614G and cell death induction more similar to D614G, indicating that, at least by these parameters, the virus did not attenuate during the course of evolution.

## Results

We isolated live virus from an individual with advanced HIV disease^15^ (defined as CD4 T cell count of less than 200 cells per microliter) who was enrolled in our longitudinal cohort to investigate the immune response to SARS-CoV-2 infection. Participants in the study cohort were sampled as soon as practicable post-diagnosis, then within the first month post-diagnosis, and subsequently at 3-month intervals^56^. In the participant with advanced HIV, continuous SARS-CoV-2 infection was detected by qPCR from combined nasopharyngeal and oropharyngeal swabs for a period of 6 months from the date of diagnosis (Figure 1A), with virus isolation performed from day 6 post-diagnosis (isolate designated D6), the first available sample for isolation, up to day 190 post-diagnosis (designated D190). We have previously sequenced these viral isolates and tested them for escape from neutralizing antibodies elicited by SARS-CoV-2 infection^15^. D6 had low to moderate immune escape from plasma sampled from convalescent individuals previously infected with either ancestral SARS-CoV-2, the Beta, or the Delta variant. However, D190 showed more extensive immune escape from ancestral virus and Delta variant infection elicited neutralizing antibodies (Figure 1B, modified analysis with data from ref^15^). Phylogenetic analysis of the infection showed a pattern consistent with the evolution of one ancestral virus infection (Figure 1C). Viral isolates D6, day 20 (D20), day 34 (D34), day 106 (D106) and D190 showed mutations in spike and other viral genes relative to ancestral virus, with neutralization escape mutation E484K along with multiple other mutations already present in D6 and the neutralization escape mutations K417T and F490S^57^ being present in the D190 isolate (Figure 1D).

**Figure 1:**
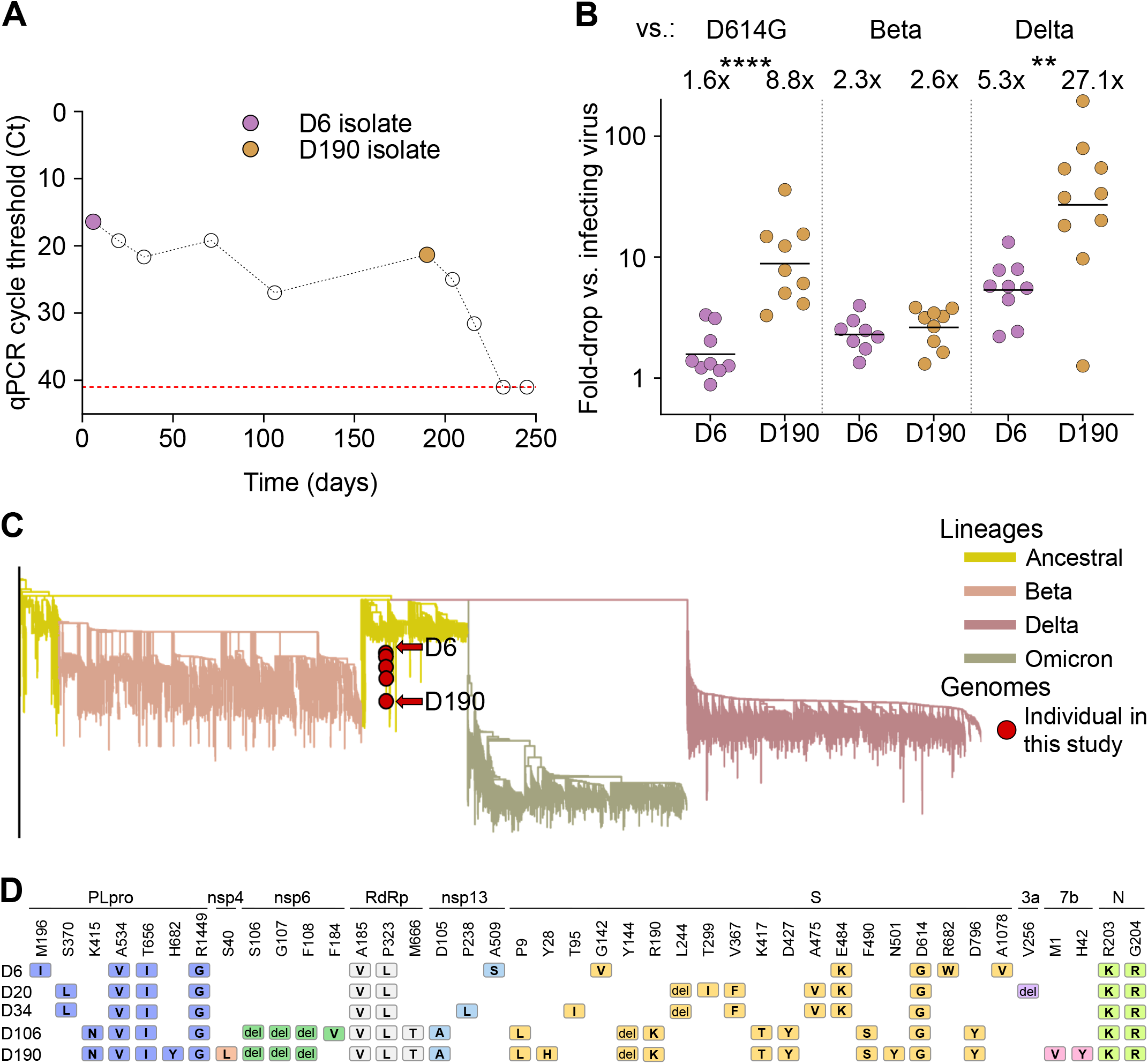
Evolution of SARS-CoV-2 from ancestral virus in an immunosuppressed individual. (**A**) SARS-CoV-2 qPCR cycle threshold (Ct) values over time. Each point represents a study visit and visits at which D6 and D190 viruses were isolated are marked with purple and orange points, respectively. Horizontal red line denotes threshold of detection. (**B**) Neutralization of D6 and D190 isolates by convalescent participant plasma with immunity elicited by D614G (left), Beta (middle) or Delta (right) variant infection. Per participant fold-drop was calculated by dividing infecting virus neutralization, quantified as FRNT_50_, by D6 or D190 neutralization. Bars represent geometric means of fold-change per participant group. p-values were **p=0.003, ****p<0.0001 by the Wilcoxon Rank Sum test. (**C**) Phylogenetic analysis of the infection, with isolates from the evolving infection tested in this study shown in red. (**D**) Substitutions and deletions in D6, D20, D34, D106, and D190 isolates relative to ancestral virus.

To test whether the virus has evolved other changes in addition to neutralizing antibody escape, we used the isolated viruses to infect the human H1299 lung cell line overexpressing the ACE2 receptor^58^. All results reported here are from live virus infections. This cell line has the endogenous histone H2AZ gene labelled with YFP by the insertion of the fluorophore sequence as an artificial exon into the first intron^59^, giving nuclear fluorescence. We used this nuclear fluorescence signal combined with automated image analysis to detect fused cells and cell number. We performed time-lapse microscopy with cells grown under controlled temperature and CO_2_ over 48 hours with images taken every 10 minutes. Uninfected cells grew until confluence with little evidence of cell death and cell-cell fusion (Video 1). In contrast, cells infected with an ancestral SARS-CoV-2 D614G isolate led to cell fusion, cell death and/or lack of cell division which became apparent about 12 hours post-infection (Video 2). These effects seemed less pronounced in cells infected with the Omicron BA.1 subvariant (Video 3) and in the D6 isolate from early after diagnosis (Video 4). However, the D190 isolate from 6 months later seemed to have increased fusogenicity and cytotoxic/cytostatic effects relative to BA.1 and D6 (Video 5).

We noted that cell nuclei become clustered together to form a contiguous region of fluorescence during cell fusion (Figure 2A). We used an automated image analysis pipeline (Figure 2 – figure supplement 1) to detect fused cells and cell number. Detection of fused cells was based on the observation that in the absence of fusion, individual cell nuclei are distinct even in confluent cell culture because they are separated by cellular cytoplasm. After fusion, cell nuclei are close together and form an area considerably larger than a single nucleus (Figure 2-figure supplement 1, Materials and methods).

**Figure 2:**
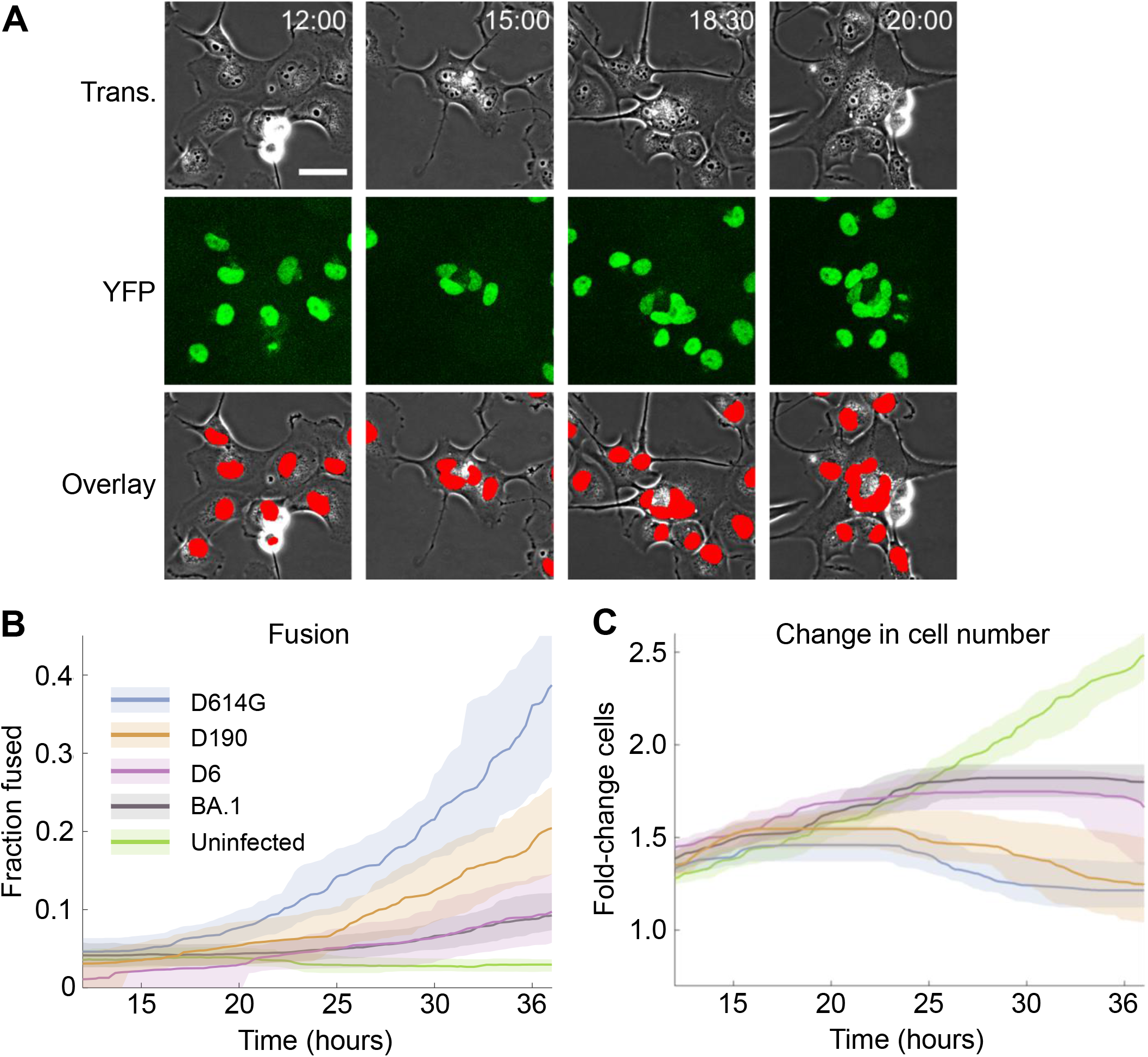
Changes in infection induced cell fusion and cell number. (**A**) Representative transmitted light (first row), nuclear YFP fluorescence (second row), and overlay images (third row) of infected cells from time-lapse imaging. Red in the overlay image denotes automated detection of nuclear area. Time of image post-infection is top right as hours:minutes. Scale bar is 50 μm. (**B**) Fraction of fused cells and (**C**) fold-change in cell number over time post-movie start, where infection is at the start of the movie. Lines and shaded areas are medians and 95% confidence intervals of 3-6 independent time-lapse experiments, with each experiment containing 12 fields of view per infection condition. Infection conditions were uninfected (green), Omicron BA.1 (grey), D6 (purple), D190 (orange) or ancestral D614G (blue) infection.

Given that our image analysis pipeline was not designed to detect dead cells, we excluded the last 12 hours of the movies when extensive cell death occurred. Quantifying fusions over multiple independent experiments showed that uninfected cell culture had a low frequency of fusions which did not increase over time. Infection by the D614G virus showed an increasing fusion frequency, with about 40% of cell nuclei in fused cells by 36 hours post-infection. The frequency of fusions was lower with BA.1 infection throughout and reached less than half of that seen in the D614G infection. The pattern in D6 virus infections was similar to BA.1, while with D190 the frequency of fusions was intermediate between BA.1 and D614G infection (Figure 2B).

To quantify changes in cell number relative to the start of the movie, we used the number of nuclei as a measure of cell number Cell numbers in uninfected cultures increased until they were about 2.5-fold higher at 36 hours relative to the start of the movie. For the D614G infection, cell numbers stopped increasing about 20 hours post-infection and started to decrease. In BA.1 and D6 infection, the number of cells also stopped increasing after about 20 hours but did not decrease to the same extent as with ancestral virus. Infection with the evolved D190 virus from late in the infection showed a similar but less pronounced decline in cell numbers as D614G (Figure 2C). Excluding nuclei in fusions from the results gave a similar pattern (Figure 2-figure supplement 2).

We used a second assay to detect cell death at 24 hours post-infection. We used this relatively early timepoint because to avoid the effect of multiple infections per cell when infection is saturating^60,61^, which happens later (see for example Video 2). Infection was detected by staining for SARS-CoV-2 nucleocapsid, and the fraction of dead infected cells determined by co-staining with a death detection dye. The positive control was addition of ethanol (Figure 3A). We compared infection by ancestral virus to BA.1 and observed that, while the fraction of infected cells was similar and slightly higher in BA.1 relative to ancestral virus, the fraction of the infected cell population staining positive for the death detection dye was lower in BA.1 (Figure 3A).

**Figure 3:**
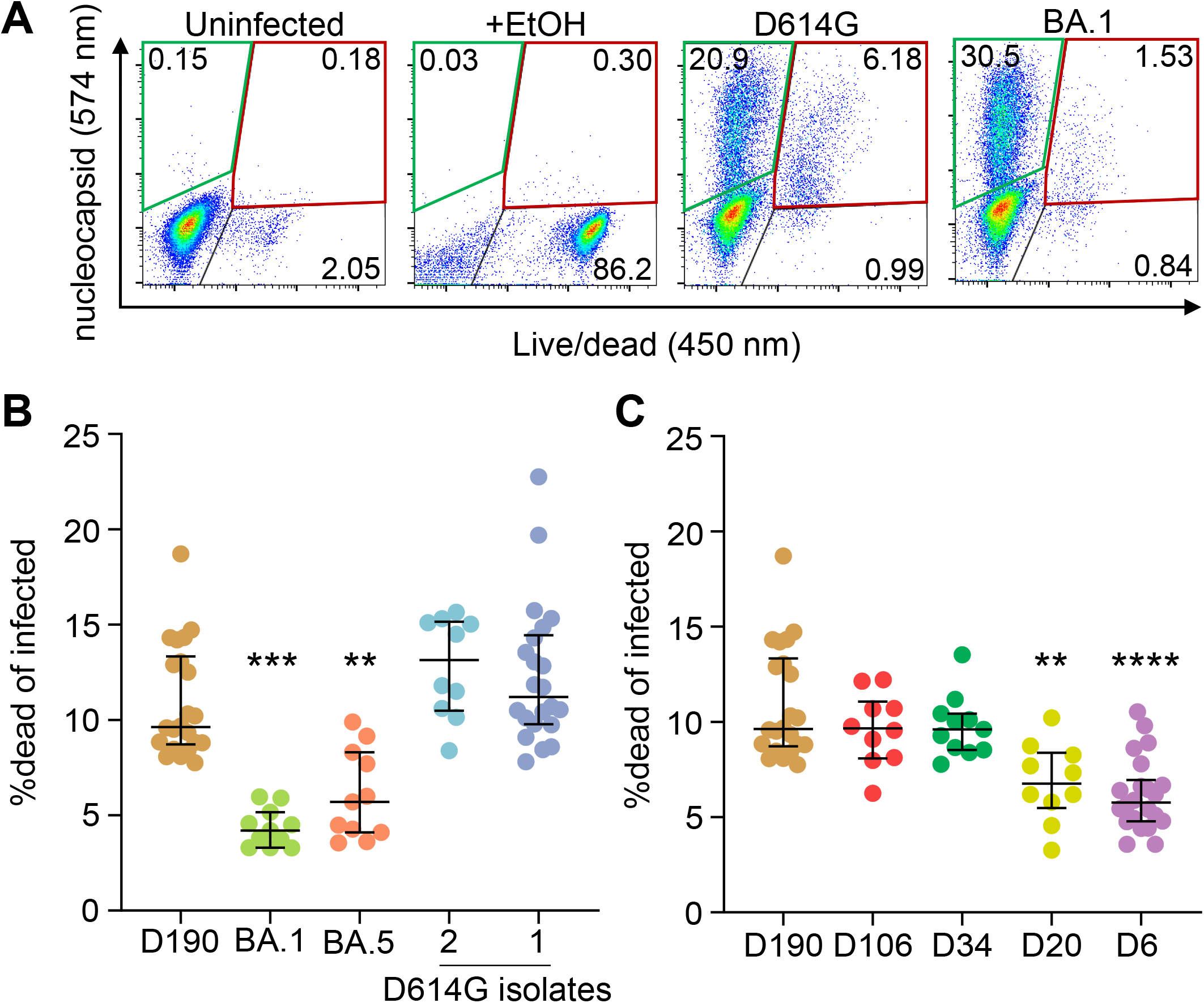
Infection by SARS-CoV-2 isolated at different times during long-term infection results in different levels of cell death. (**A**) Flow cytometry gating strategy. First panel shows uninfected cells, second panel shows 80% ethanol-treated cells (positive control), and third and fourth panels show live infected cells (green gate) and dead infected cells (red gate) 24 hours post-infection with D614G and Omicron BA.1. Numbers represent percentages of cells in the corresponding quadrants. (**B**) Fraction of dead cells 24 hours post-infection in infections by D190, Omicron subvariants BA.1 and BA.5, and two independent isolates of ancestral virus with the D614G substitution. (**C**) Fraction of dead cells 24 hours post-infection in infections by D190, D106, D34, D20, and D6. Horizontal bars represent medians with interquartile ranges of 10-22 replicates from 3-8 independent experiments with all experiments containing D190 and the D614G isolate 1 for reference. p-values were determined by the Kruskal-Wallis test with Dunn multiple comparisons correction, with all comparisons to D190. Significant p-values were ***p=0.0001 (D190 vs. BA.1), **p=0.0015 (D190 vs. BA.5), ****p< 0.0001 (D190 vs. D6) and **p=0.0064 (D190 vs. D20).

We tested two independent isolates of ancestral D614G virus (D614G.1, D614G.2, see Materials and methods), Omicron subvariants BA.1 and BA.5, the D6 and D190 isolates, and isolates from study visits at day 20 (D20), 34 (D34), and 106 (D106) post-diagnosis. We then calculated the ratio of the fraction of dead infected cells to total infected cells at 24 hours post-infection. We observed that both isolates of ancestral virus caused cell death to a similar extent, with about 11-13% of infected cells being detected as dead. The frequency of death was about 3-fold lower for BA.1, at 4.2%, and was higher for BA.5, at 5.7%. The frequency of D190 induced cell death, at 9.6%, was significantly higher than both BA.1 and BA.5 and more like the ancestral virus isolates (Figure 3B). Among the isolates from the earlier timepoints, D6 infection led to 5.6% dead infected cells, a similar frequency of cell death induction as BA.5. The frequency of cell death increased to 6.8% for D20 and to 9.6% for D34 and D106. The cell death frequency was significantly lower in D6 and D20 relative to D190 infections (Figure 3C).

Because cell-cell fusion depends on cell surface spike expression, we investigated surface spike expression on the infected cells in infection foci (where each focus contains on the order of 10 cells, see^31^) formed by live virus on a confluent cell monolayer 18 hours post-infection. To ensure that we did not inadvertently permeabilize the cells as part of the staining procedure, thereby including intracellular spike, we also stained for SARS-CoV-2 nucleocapsid protein, not present on the cell surface (Figure 4A). We observed that in unpermeabilized cells, surface spike was readily detected. In contrast, nucleocapsid protein was not detected on the cell surface, although levels were high after permeabilization. We then measured mean intensity, which is independent of focus size, and normalized by the mean intensity of the ancestral SARS-CoV-2 isolate (D614G isolate 1) included in each of the independent experiments. We observed that the two ancestral virus isolates were similar in spike expression. BA.1 infection showed significantly reduced cell surface spike, but levels increased moderately in BA.5 infection. D190 infection had significantly lower spike than the ancestral virus isolates but higher surface spike relative to D6 infection (Figure 4B).

**Figure 4:**
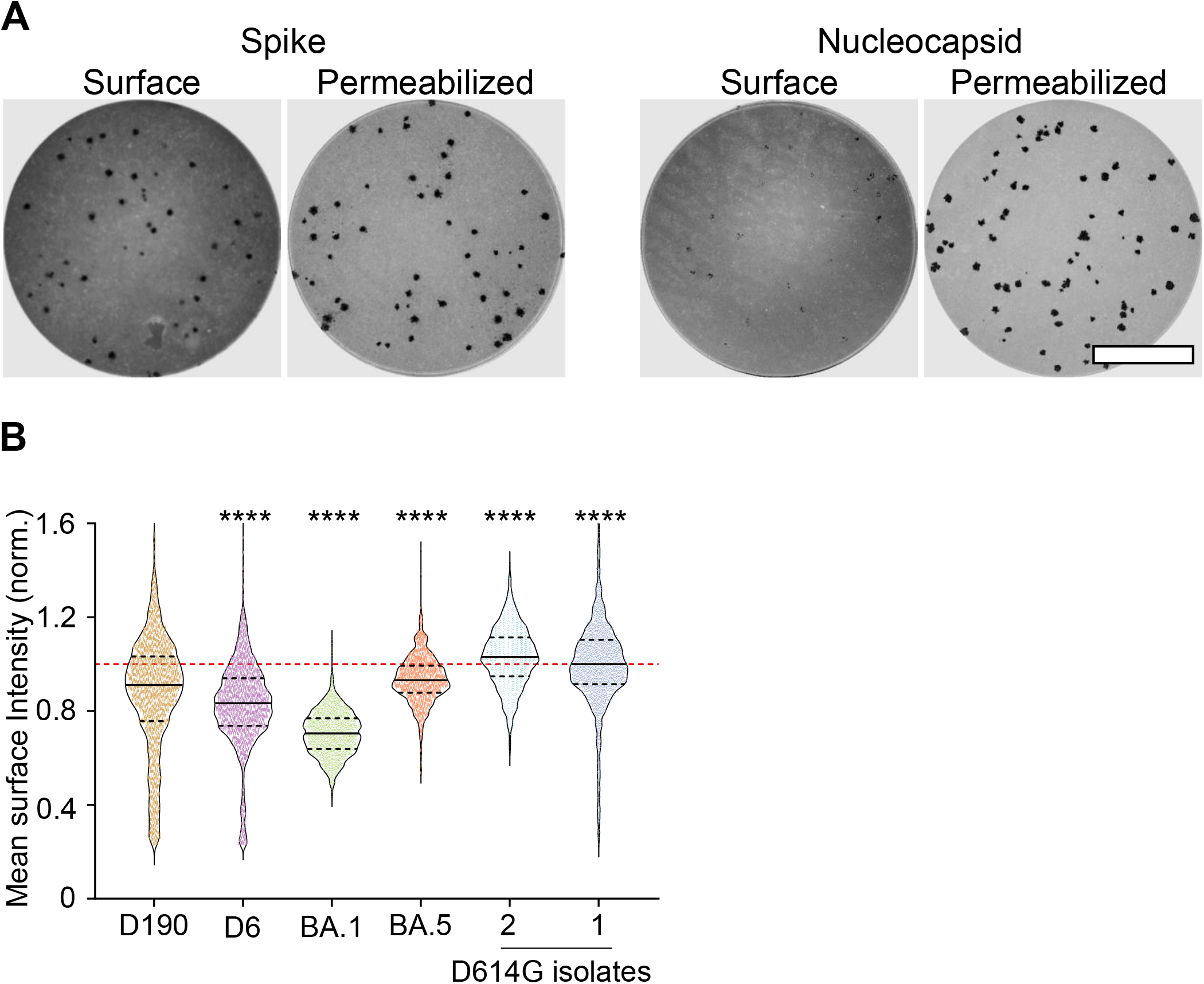
Changes in spike cell surface expression during evolution. (**A**) Expression of spike (left 2 wells) and nucleocapsid (right 2 wells) in representative infection foci formed by ancestral virus at 18 hours post-infection. For each set of two images, the cells in the left image were stained without permeabilization and show cell surface expression of either spike or nucleocapsid. The cells on the right have been permeabilized and show both surface and intracellular spike or nucleocapsid expression. Each image represents the complete area of one well of a 96-well plate in a live virus focus forming assay. Bar is 2 mm. (**B**) The mean intensity of surface spike levels of individual foci, normalized per experiment to the mean of the ancestral virus cell surface spike expression obtained in the experiment. Mean intensity data from 2038 (D190), 1913 (D6), 1627 (BA.1), 1237 (BA.5), 1028 (D614G isolate 2), and 3052 (D614G isolate 1) foci from 3-6 independent experiments. p-values were determined by the Kruskal-Wallis test with Dunn multiple comparisons correction, with all comparisons to D190 and all ****p<0.0001.

## Discussion

Here we investigated the effect of evolutionary changes which happened in SARS-CoV-2 in prolonged infection in an individual immunosuppressed because of advanced HIV disease. We assayed a virus isolated from early in the course of infection at day 6 post-diagnosis, at day 190 post-diagnosis, as well as at the intermediate timepoints. The day 6 isolate already evolved a considerable number of mutations, or alternatively, the infecting virus was mutated. Between day 6 and day 190 post-diagnosis, the infection evolved immune escape from neutralizing antibodies, consistent with what is observed in long-term SARS-CoV-2 infections in individuals with immunosuppression^24^. However, the ability of the virus to induce cell fusion and cell death upon infection also changed through the course of evolution. While the early viral isolate induced lower cell fusion and death compared to ancestral virus, induction of fusion and cell death increased with evolution

We cannot determine whether the infecting virus evolved a reduced ability to cause cell fusion and death by day 6 post-diagnosis, or whether this was a property of the infecting virus. Symptom onset for the infection was reported to be 10 days pre-diagnosis^16^, which a seems a short time to evolve substantial differences relative to the ancestral virus, but we do not know for certain how long the individual was infected. In addition, SARS-CoV-2 has been documented to evolve relatively rapidly in some cases^62^. However, we have determined that the subsequently isolated viruses evolved an enhanced ability to fuse cells and cause cell death relative to the early isolate.

Consistent with the notion that highly mutated variants may not all have attenuated pathogenicity, Beta variant infections in South Africa did not show lower pathogenicity relative to ancestral virus infections^56^. This does not imply that there is no selective advantage in attenuation. For example, variants which do not lead to more severe disease may be more transmissible since the infected individual may continue daily activities and therefore transmit to more people.

A possible selective pressure that results in the evolution of increased cell fusion is escape from neutralizing antibodies. We and others have observed that cell-cell spread of SARS-CoV-2 using this mechanism is insensitive to neutralizing antibodies, although inhibition is possible at high antibody concentrations^43,63-65^. The immunosuppressed participant from whom the D6 and D190 viruses were isolated showed increasing neutralizing antibody activity during the infection^15^, and development of enhanced cell-cell spread may have been selected for to escape neutralization. Higher infection elicited cell death may be related to increased fusogenicity. This relationship seems to be present in the time-lapse data and is cell death, and cell death because of syncytium formation is well characterized for HIV^66^.

The Omicron subvariants circulating since November 2021 have led to infections with a lower probability of developing severe disease^1,2,4,67^. Moreover, Omicron infection boosts the immune response against other variants as well as against other Omicron subvariants^68^, likely maintaining protective immunity through infection. The results presented here, with the limitations that we tested cellular parameters as measures of pathogenicity and viruses from only one case of long-term evolution, may indicate that SARS-CoV-2 evolution in long-term infection does not have to result in attenuation. It may indicate that a future variant could be more pathogenic than currently circulating Omicron strains.

## Materials and methods

### Informed consent and ethical statement

Swabs for the isolation of ancestral (D614G.1 and D614G.2), Beta, Delta, D6, D20, D34, D106, and D190 viruses and blood samples to test virus neutralization were obtained after written informed consent from adults with PCR-confirmed SARS-CoV-2 infection who were enrolled in a prospective cohort study at the Africa Health Research Institute approved by the Biomedical Research Ethics Committee at the University of KwaZulu–Natal (reference BREC/00001275/2020). The Omicron/BA.1 was isolated from a residual swab sample with SARS-CoV-2 isolation from the sample approved by the University of the Witwatersrand Human Research Ethics Committee (HREC) (ref. M210752). The sample to isolate Omicron/BA.5 was collected after written informed consent as part of the COVID-19 transmission and natural history in KwaZulu-Natal, South Africa: Epidemiological Investigation to Guide Prevention and Clinical Care in the Centre for the AIDS Programme of Research in South Africa (CAPRISA) study and approved by the Biomedical Research Ethics Committee at the University of KwaZulu–Natal (reference BREC/00001195/2020, BREC/00003106/2021).

### Reagent availability statement

Isolates and raw image files are available upon reasonable request. Sequences of isolated SARS-CoV-2 used in this study have been deposited in GISAID with accession:

| Virus | GISAID number | PANGO |
| --- | --- | --- |
| D614G.1 | EPI_ISL_602626.1 | B.1 |
| D614G.2 | EPI_ISL_602622 | B.1.1.117 |
| BA.1 | EPI_ISL_7886688 | BA.1 |
| BA.5 | EPI_ISL_12268493.2 | BA.5 |
| 0027-D6 | EPI_ISL_15541746 | B.1.1.273 |
| 0027-D20 | EPI_ISL_15541747 | B.1.1.273 |
| 0027-D34 | EPI_ISL_15541748 | B.1.1.273 |
| 0027-D106 | EPI_ISL_15541750 | B.1.1.273 |
| 0027-D190 | EPI_ISL_2397313 | B.1.1.273 |

### Whole-genome sequencing, genome assembly and phylogenetic analysis

RNA was extracted on an automated Chemagic 360 instrument, using the CMG-1049 kit (Perkin Elmer, Hamburg, Germany). The RNA was stored at −80°C prior to use. Libraries for whole genome sequencing were prepared using either the Oxford Nanopore Midnight protocol with Rapid Barcoding or the Illumina COVIDseq Assay. For the Illumina COVIDseq assay, the libraries were prepared according to the manufacturer’s protocol. Briefly, amplicons were tagmented, followed by indexing using the Nextera UD Indexes Set A. Sequencing libraries were pooled, normalized to 4 nM and denatured with 0.2 N sodium acetate. An 8 pM sample library was spiked with 1% PhiX (PhiX Control v3 adaptor-ligated library used as a control). We sequenced libraries on a 500-cycle v2 MiSeq Reagent Kit on the Illumina MiSeq instrument (Illumina). On the Illumina NextSeq 550 instrument, sequencing was performed using the Illumina COVIDSeq protocol (Illumina Inc, USA), an amplicon-based next-generation sequencing approach. The first strand synthesis was carried using random hexamers primers from Illumina and the synthesized cDNA underwent two separate multiplex PCR reactions. The pooled PCR amplified products were processed for tagmentation and adapter ligation using IDT for Illumina Nextera UD Indexes. Further enrichment and cleanup was performed as per protocols provided by the manufacturer (Illumina Inc). Pooled samples were quantified using Qubit 3.0 or 4.0 fluorometer (Invitrogen Inc.) using the Qubit dsDNA High Sensitivity assay according to manufacturer’s instructions. The fragment sizes were analyzed using TapeStation 4200 (Invitrogen). The pooled libraries were further normalized to 4nM concentration and 25 μL of each normalized pool containing unique index adapter sets were combined in a new tube. The final library pool was denatured and neutralized with 0.2N sodium hydroxide and 200 mM Tris-HCL (pH7), respectively. 1.5 pM sample library was spiked with 2% PhiX. Libraries were loaded onto a 300-cycle NextSeq 500/550 HighOutput Kit v2 and run on the Illumina NextSeq 550 instrument (Illumina, San Diego, CA, USA). For Oxford Nanopore sequencing, the Midnight primer kit was used as described by Freed and Silander55. cDNA synthesis was performed on the extracted RNA using LunaScript RT mastermix (New England BioLabs) followed by gene-specific multiplex PCR using the Midnight Primer pools which produce 1200bp amplicons which overlap to cover the 30-kb SARS-CoV-2 genome. Amplicons from each pool were pooled and used neat for barcoding with the Oxford Nanopore Rapid Barcoding kit as per the manufacturer’s protocol. Barcoded samples were pooled and bead-purified. After the bead clean-up, the library was loaded on a prepared R9.4.1 flow-cell. A GridION X5 or MinION sequencing run was initiated using MinKNOW software with the base-call setting switched off. We assembled paired-end and nanopore.fastq reads using Genome Detective 1.132 (https://www.genomedetective.com) which was updated for the accurate assembly and variant calling of tiled primer amplicon Illumina or Oxford Nanopore reads, and the Coronavirus Typing Tool56. For Illumina assembly, GATK HaploTypeCaller -- min-pruning 0 argument was added to increase mutation calling sensitivity near sequencing gaps. For Nanopore, low coverage regions with poor alignment quality (<85% variant homogeneity) near sequencing/amplicon ends were masked to be robust against primer drop-out experienced in the Spike gene, and the sensitivity for detecting short inserts using a region-local global alignment of reads, was increased. In addition, we also used the wf_artic (ARTIC SARS-CoV-2) pipeline as built using the nextflow workflow framework57. In some instances, mutations were confirmed visually with .bam files using Geneious software V2020.1.2 (Biomatters). The reference genome used throughout the assembly process was NC_045512.2 (numbering equivalent to MN908947.3). For lineage classification, we used the widespread dynamic lineage classification method from the ‘Phylogenetic Assignment of Named Global Outbreak Lineages’ (PANGOLIN) software suite (https://github.com/hCoV-2019/pangolin).

### Cells

The H1299-E3 (H1299-ACE2, clone E3, H1299 originally from ATCC as CRL-5803) cell line was derived from H1299 as described in our previous work^58,69^. The H1299-E3 cells were propagated in growth medium consisting of complete Roswell Park Memorial Institute (RPMI) 1640 medium with 10% fetal bovine serum (Hyclone) containing 10mM of hydroxyethylpiperazine ethanesulfonic acid (HEPES), 1mM sodium pyruvate, 2mM L-glutamine and 0.1mM nonessential amino acids (all Sigma-Aldrich). Cells were passaged every second day. For virus isolation, Vero E6 cells (originally ATCC CRL-1586, obtained from Cellonex in South Africa) were propagated in complete growth medium consisting of Dulbecco’s Modified Eagle Medium (DMEM) with 10% fetal bovine serum (Hyclone) containing 10mM of HEPES, 1mM sodium pyruvate, 2mM L-glutamine and 0.1mM nonessential amino acids (all Sigma-Aldrich). Vero E6 cells were passaged every 3–4 days.

### Virus isolation

All work with live virus was performed in Biosafety Level 3 containment using protocols for SARS-CoV-2 approved by the Africa Health Research Institute Biosafety Committee. ACE2-expressing H1299-E3 cells were seeded at 4.5 × 10^5^ cells in a 6 well plate well and incubated for 18–20 h. After one Dulbecco’s phosphate-buffered saline (DPBS) wash, the sub-confluent cell monolayer was inoculated with 500 μL universal transport medium from swabs diluted 1:1 with growth medium filtered through a 0.45-μm filter. Cells were incubated for 1 h. Wells were then filled with 3 mL complete growth medium. After 4 days of infection (completion of passage 1 (P1)), cells were trypsinized (Sigma-Aldrich), centrifuged at 300 rcf for 3 min and resuspended in 4 mL growth medium. Then all infected cells were added to Vero E6 cells that had been seeded at 1.5 × 10^5^ cells per mL, 20 mL total, 18–20 h earlier in a T75 flask for cell-to-cell infection. The coculture of ACE2-expressing H1299-E3 and Vero E6 cells was incubated for 1 h and the flask was filled with 20 mL of complete growth medium and incubated for 4 days. The viral supernatant from this culture (passage 2 (P2) stock) was used for experiments.

### Time-lapse microscopy

6-well glass bottom plates (MatTek) were coated with 300 μL of 0.001% fibronectin (Sigma-Aldrich) in DPBS-/-(Gibco), incubated for 90 mins, then washed 3x with DPBS. H1299-E3 cells were then immediately plated at 60,000 cells per coated well. The next day the cells were infected at 1000 focus-forming units in 1 mL growth media per well. Cell–virus mixtures were incubated for 1 h at 37 °C, 5% CO_2_ then an additional 1 mL of growth media was added. Infections were imaged using a Metamorph controlled Nikon TiE motorized microscope (Nikon Corporation) in a Biosafety Level 3 Facility with a 20x, 0.75 NA phase objective. Images were captured using an 888 EMCCD camera (Andor). Temperature (37°C), humidity and CO_2_ (5%) were controlled using an environmental chamber (OKO Labs). Excitation source was 488 laser line and emission was detected through a Semrock Brightline quad band 440–40 /521-21/607-34/700-45 nm filter. For each well, 12 randomly chosen fields of view were captured every 10 minutes.

### Image analysis

Timelapse microscopy images were analysed using custom MATLAB v.2019b (MathWorks) script and using the MATLAB Image Analysis Toolbox. For each frame in the movie, both the transmitted light and fluorescent images were used. A coarse segmentation was first performed using the transmitted light image of the cells. Images were flatfield corrected and contrast was enhanced by setting the top and bottom 1% of all pixel intensities to 1 and 0 respectively. The built-in function “rangefilt” was used to determine areas of high contrast (where high pixel intensities were immediately adjacent to low pixel intensities) in the image which corresponded to cell borders. Processed images were also median filtered and holes to filled within segmented objects. The image was then thresholded to remove background signal and obtain a mask of areas occupied by cells. The mask generated from the transmitted light image was then used to remove objects that were not in areas occupied by cells. In the fluorescence images corresponding to the transmitted light images. Fluorescence images were then processed using flatfield correction, contrast enhancement and median filtering. Fluorescent cell nuclei in the YFP channel were used to generate a binary mask for contiguous objects in each image after thresholding. Each object was categorized as multi-nucleate or uni-nucleate based on pixel area, where the threshold for a single nucleus was calculated as the mean area of objects/nuclei in the uninfected condition, at 12 hours post-movie start, + 3 standard deviations of the mean. The fraction of cells in fusions was calculated by dividing the total pixel area of objects above the single nucleus threshold by the total pixel area occupied by nuclei in the same frame. The number of nuclei was calculated by dividing the total pixel area occupied by nuclei by the mean pixel area of one nucleus in the uninfected condition at 12 hours post-movie start.

### Detection of infected cell death

H1299-E3 cells were plated at 60,000 cells per well in 6-well plates (Corning) 1 day pre-infection. The next day the cells were infected at 1000 focus-forming units in 1 mL growth media per well. Cell–virus mixtures were incubated for 1 h at 37 °C, 5% CO_2_ then an additional 1 mL of growth media was added. 24 hours post-infection, cells were trypsinised (Sigma-Aldrich), collected and stained with Blue Live/Dead stain as per manufacturer instructions (L34961, ThermosScientific). The samples were then washed in 1 mL PBS-/- and resuspended in Cytofix/Cytoperm (BD Biosciences) for 20 min at 4°C in the dark. The samples were then stained with 0.5 μg/mL anti-SARS-CoV-2 nucleocapsid-PE (ab283244, Abcam) for 1 hour at 4°C in the dark. Cells were analysed on an Aria Fusion (BD). Data was analysed using FlowJo and Graphpad Prism 9.4.1 software.

### Staining for cell-surface and total spike in a focus forming assay

H1299-E3 cells were plated in a 96-well plate (Corning) at 20,000 cells per well 1 day pre-infection. Virus stocks were used at 100 focus-forming units per microwell. Cells were infected with 100 μL of the virus for 1 h, then 100 μL of a 1X RPMI 1640 (Sigma-Aldrich, R6504), 1.5% carboxymethylcellulose (Sigma-Aldrich, C4888) overlay was added without removing the inoculum. Cells were fixed at 18 hours post-infection using 4% methanol-free formaldehyde (ThermoScientific) for 20 minutes. For staining of foci, a rabbit anti-SARS-CoV-2 spike monoclonal antibody (mAb BS-R2B12, GenScript A02058) at 0.5 ug/mL or a rabbit anti-SARS-CoV-2 nucleocapsid monoclonal (ab271180 Abcam) at 1 ug/mL were used as the primary detection antibody. Antibody was resuspended in either a permeabilization buffer containing 0.1% saponin (Sigma-Aldrich), 0.1% BSA (Sigma-Aldrich), and 0.05% Tween-20 (Sigma-Aldrich) in PBS+/+ or a non-permeabilization buffer containing 0.1% BSA and 0.05% Tween-20 in PBS+/+. Plates were incubated with primary antibody at room temperature for 2 hr with shaking, then washed with wash buffer containing 0.05% tween in PBS+/+. Secondary goat anti-rabbit horseradish peroxidase (Abcam ab205718) was added at 1 ug/mL in either permeabilization or non-permeabilization buffers as described above and incubated for 2 hours at room temperature with shaking. TrueBlue peroxidase substrate (SeraCare 5510-0030) was then added at 50uL per well and incubated for 15 minutes at room temperature. Plates were washed in distilled water and then dried for 2 hours and imaged in an ImmunoSpot Ultra-V S6-02-6140 Analyzer ELISPOT instrument with BioSpot Professional built-in image analysis (C.T.L). Data was analyzed using Graphpad Prism 9.4.1.

### Statistics and fitting

Fitting was performed using MATLAB v.2019b. Neutralization data were fit to:

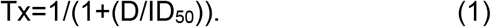

Here Tx is the number of foci normalized to the number of foci in the absence of plasma on the same plate at dilution D and ID_50_ is the plasma dilution giving 50% neutralization. FRNT_50_ = 1/ID_50_. Values of FRNT_50_ <1 are set to 1 (undiluted), the lowest measurable value. We note that the most concentrated plasma dilution was 1:25 and therefore FRNT_50_ < 25 were extrapolated. Fold-change was calculated by dividing the FRNT_50_ obtained for the homologous virus (the virus which elicited the immunity, e.g. D614G) by the FRNT_50_ heterologous virus (e.g. D190) per participant, then calculating the geometric mean and 95% confidence intervals over all participant values. The 95% confidence intervals on the median in the time-lapse microscopy data was calculated by first ranking the values in ascending order, then finding the ranks of the lower and upper confidence interval by:

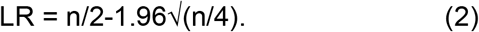

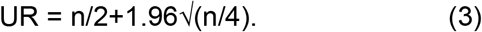

Here LR is lower 95% interval index in the ranked vector, UR is the upper 95% interval index in the ranked vector. Values at indexes LR and UR were the lower 95% and upper 95% confidence intervals, respectively. Other statistical tests, measures of central tendency and confidence intervals were performed in GraphPad Prism version 9.4.1.

## Data Availability

Isolates and raw image files are available upon reasonable request. Sequences of isolated SARS-CoV-2 used in this study have been deposited in GISAID.

https://gisaid.org/

## Acknowledgements

This study was supported by the Bill and Melinda Gates award INV-018944 (AS), National Institutes of Health award R01 AI138546 (AS), and South African Medical Research Council Award 6084COAP2020 (AS). The funders had no role in study design, data collection and analysis, decision to publish, or preparation of the manuscript.

## Competing interest statement

AS received an honorarium for a talk given to Pfizer employees.

## Figure legends

**Figure 2 – figure supplement 1:**
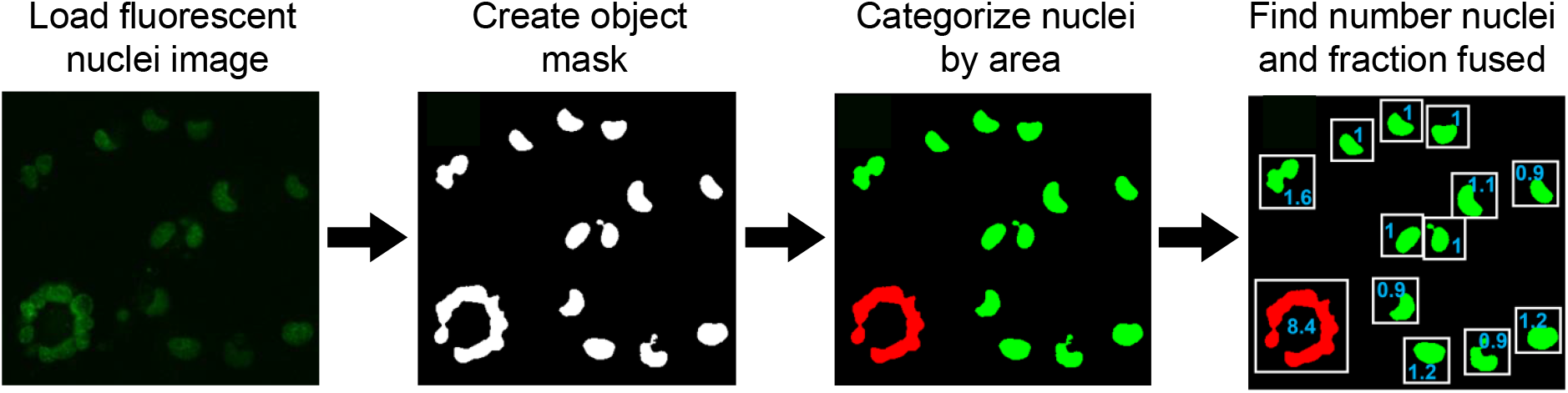
Overview of automated image analysis. Images of fluorescently labelled nuclei were loaded to MATLAB 2019b and a binary mask was generated by thresholding on the fluorescent signal to separate individual objects. An object was classified as multinucleate/fused (red) if the object’s area was larger than the maximum determined size for a single nucleus, and uni-nucleate (green) otherwise. The maximum size threshold for a single nucleus was calculated as the mean nuclear area of uninfected cells at 12 hours post-movie start plus 3 standard deviations of the mean. The total number of nuclei per movie frame was determined by dividing the sum of the nuclear area by the mean area of a single nucleus, and the fraction of fused cells was determined by dividing the sum of the area of nuclei scored as fused by the sum of the total nuclear area.

**Figure 2 – figure supplement 2:**
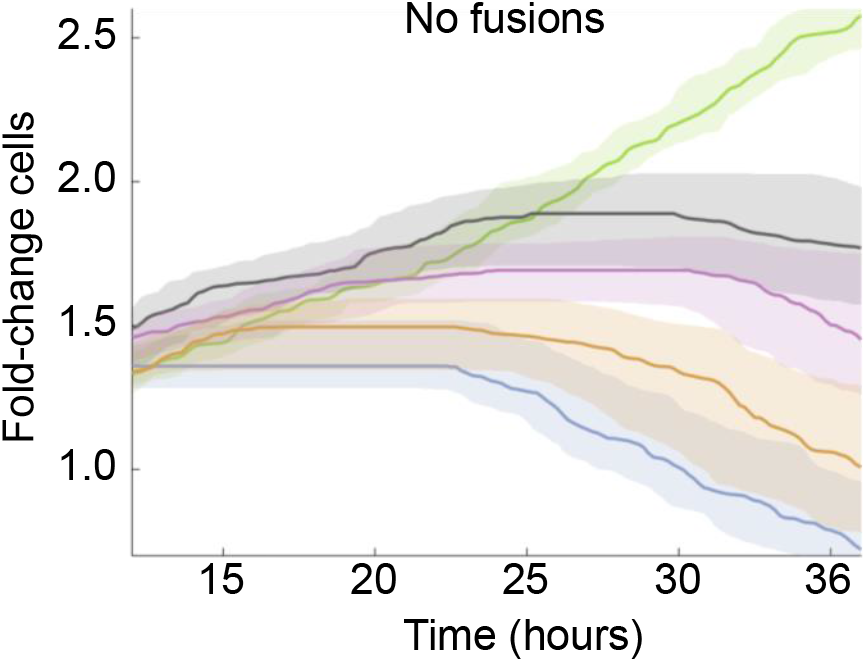
Fold-change in cell number post-infection with exclusion of fused cells. Fold-change in cell number over time post-movie start. Total number of unfused cells was determined by subtracting the total area of fused nuclei from the sum of the total area occupied by nuclei, then dividing the remaining nuclear area by the mean area of a single nucleus. Lines and shaded areas are median and 95% confidence intervals of 3-6 independent time-lapse experiments containing 12 fields of view each per infection condition. Infection conditions were uninfected (green), BA.1 (grey), D6 (purple), D190 (orange) or ancestral D614G (blue).

## Video captions

**Video 1:** Representative field of view of uninfected H1299-ACE2 cells over 50 hours of imaging at 10-minute intervals between frames.

**Video 2:** Representative field of view of ancestral D614G virus infected H1299-ACE2 cells over 50 hours of imaging at 10-minute intervals between frames.

**Video 3:** Representative field of view of Omicron BA.1 virus infected H1299-ACE2 cells over 50 hours of imaging at 10-minute intervals between frames.

**Video 4:** Representative field of view of D6 isolate virus infected H1299-ACE2 cells over 50 hours of imaging at 10-minute intervals between frames.

**Video 5:** Representative field of view of D190 isolate virus infected H1299-ACE2 cells over 50 hours of imaging at 10-minute intervals between frames.

## Notes

### Author Declarations

Swabs for the isolation of ancestral (D614G.1 and D614G.2), Beta, Delta, D6, D20, D34, D106, and D190 viruses and blood samples to test virus neutralization were obtained after written informed consent from adults with PCR-confirmed SARS-CoV-2 infection who were enrolled in a prospective cohort study at the Africa Health Research Institute approved by the Biomedical Research Ethics Committee at the University of KwaZulu-Natal (reference BREC/00001275/2020). The Omicron/BA.1 was isolated from a residual swab sample with SARS-CoV-2 isolation from the sample approved by the University of the Witwatersrand Human Research Ethics Committee (HREC) (ref. M210752). The sample to isolate Omicron/BA.5 was collected after written informed consent as part of the COVID-19 transmission and natural history in KwaZulu-Natal, South Africa: Epidemiological Investigation to Guide Prevention and Clinical Care in the Centre for the AIDS Programme of Research in South Africa (CAPRISA) study and approved by the Biomedical Research Ethics Committee at the University of KwaZulu-Natal (reference BREC/00001195/2020, BREC/00003106/2021).

### Summary of Updates

Added discussion on possible reasons for the evolution of increased fusogenicity

## References

1 Sigal, A., Milo, R. & Jassat, W. Estimating disease severity of Omicron and Delta SARS-CoV-2 infections. Nat Rev Immunol 22, 267–269 (2022). PMC9002222. 10.1038/s41577-022-00720-5

2 Sigal, A. Milder disease with Omicron: is it the virus or the pre-existing immunity? Nature Reviews Immunology 22, 69–71 (2022). 10.1038/s41577-022-00678-4

3 Whitaker, M., Elliott, J., Bodinier, B., Barclay, W., Ward, H., Cooke, G., Donnelly, C. A., Chadeau-Hyam, M. & Elliott, P. Variant-specific symptoms of COVID-19 in a study of 1,542,510 adults in England. Nat Commun 13, 6856 (2022). PMC9651890. 10.1038/s41467-022-34244-2

4 Ward, I. L., Bermingham, C., Ayoubkhani, D., Gethings, O. J., Pouwels, K. B., Yates, T., Khunti, K., Hippisley-Cox, J., Banerjee, A., Walker, A. S. & Nafilyan, V. Risk of covid-19 related deaths for SARS-CoV-2 omicron (B.1.1.529) compared with delta (B.1.617.2): retrospective cohort study. Bmj 378, e070695 (2022). PMC9344192. 10.1136/bmj-2022-070695

5 Griffin, B. D., Chan, M., Tailor, N., Mendoza, E. J., Leung, A., Warner, B. M., Duggan, A. T., Moffat, E., He, S., Garnett, L., Tran, K. N., Banadyga, L., Albietz, A., Tierney, K., Audet, J., Bello, A., Vendramelli, R., Boese, A. S., Fernando, L., Lindsay, L. R., Jardine, C. M., Wood, H., Poliquin, G., Strong, J. E., Drebot, M., Safronetz, D., Embury-Hyatt, C. & Kobasa, D. SARS-CoV-2 infection and transmission in the North American deer mouse. Nat Commun 12, 3612 (2021). PMC8203675. 10.1038/s41467-021-23848-9

6 Meekins, D. A., Gaudreault, N. N. & Richt, J. A. Natural and Experimental SARS-CoV-2 Infection in Domestic and Wild Animals. Viruses 13 (2021). PMC8540328. 10.3390/v13101993

7 Kuchipudi, S. V., Surendran-Nair, M., Ruden, R. M., Yon, M., Nissly, R. H., Vandegrift, K. J., Nelli, R. K., Li, L., Jayarao, B. M., Maranas, C. D., Levine, N., Willgert, K., Conlan, A. J. K., Olsen, R. J., Davis, J. J., Musser, J. M., Hudson, P. J. & Kapur, V. Multiple spillovers from humans and onward transmission of SARS-CoV-2 in white-tailed deer. Proc Natl Acad Sci U S A 119 (2022). PMC8833191. 10.1073/pnas.2121644119

8 Hale, V. L., Dennis, P. M., McBride, D. S., Nolting, J. M., Madden, C., Huey, D., Ehrlich, M., Grieser, J., Winston, J., Lombardi, D., Gibson, S., Saif, L., Killian, M. L., Lantz, K., Tell, R. M., Torchetti, M., Robbe-Austerman, S., Nelson, M. I., Faith, S. A. & Bowman, A. S. SARS-CoV-2 infection in free-ranging white-tailed deer. Nature 602, 481–486 (2022). PMC8857059. 10.1038/s41586-021-04353-x

9 Koeppel, K. N., Mendes, A., Strydom, A., Rotherham, L., Mulumba, M. & Venter, M. SARS-CoV-2 Reverse Zoonoses to Pumas and Lions, South Africa. Viruses 14 (2022). PMC8778549. 10.3390/v14010120

10 Fenollar, F., Mediannikov, O., Maurin, M., Devaux, C., Colson, P., Levasseur, A., Fournier, P. E. & Raoult, D. Mink, SARS-CoV-2, and the Human-Animal Interface. Front Microbiol 12, 663815 (2021). PMC8047314. 10.3389/fmicb.2021.663815

11 Bayarri-Olmos, R., Rosbjerg, A., Johnsen, L. B., Helgstrand, C., Bak-Thomsen, T., Garred, P. & Skjoedt, M. O. The SARS-CoV-2 Y453F mink variant displays a pronounced increase in ACE-2 affinity but does not challenge antibody neutralization. J Biol Chem 296, 100536 (2021). PMC7948531. 10.1016/j.jbc.2021.100536

12 Hoffmann, M., Zhang, L., Krüger, N., Graichen, L., Kleine-Weber, H., Hofmann-Winkler, H., Kempf, A., Nessler, S., Riggert, J., Winkler, M. S., Schulz, S., Jäck, H. M. & Pöhlmann, S. SARS-CoV-2 mutations acquired in mink reduce antibody-mediated neutralization. Cell Rep 35, 109017 (2021). PMC8018833. 10.1016/j.celrep.2021.109017

13 Ren, W., Lan, J., Ju, X., Gong, M., Long, Q., Zhu, Z., Yu, Y., Wu, J., Zhong, J., Zhang, R., Fan, S., Zhong, G., Huang, A., Wang, X. & Ding, Q. Mutation Y453F in the spike protein of SARS-CoV-2 enhances interaction with the mink ACE2 receptor for host adaption. PLoS Pathog 17, e1010053 (2021). PMC8601601. 10.1371/journal.ppat.1010053

14 Zhou, J., Peacock, T. P., Brown, J. C., Goldhill, D. H., Elrefaey, A. M. E., Penrice-Randal, R., Cowton, V. M., De Lorenzo, G., Furnon, W., Harvey, W. T., Kugathasan, R., Frise, R., Baillon, L., Lassaunière, R., Thakur, N., Gallo, G., Goldswain, H., Donovan-Banfield, I., Dong, X., Randle, N. P., Sweeney, F., Glynn, M. C., Quantrill, J. L., McKay, P. F., Patel, A. H., Palmarini, M., Hiscox, J. A., Bailey, D. & Barclay, W. S. Mutations that adapt SARS-CoV-2 to mink or ferret do not increase fitness in the human airway. Cell Rep 38, 110344 (2022). PMC8768428. 10.1016/j.celrep.2022.110344

15 Cele, S., Karim, F., Lustig, G., San, J. E., Hermanus, T., Tegally, H., Snyman, J., Moyo-Gwete, T., Wilkinson, E., Bernstein, M., Khan, K., Hwa, S.-H., Tilles, S. W., Singh, L., Giandhari, J., Mthabela, N., Mazibuko, M., Ganga, Y., Gosnell, B. I., Abdool Karim, S. S., Hanekom, W., Van Voorhis, W. C., Ndung’u, T., Lessells, R. J., Moore, P. L., Moosa, M.-Y. S., de Oliveira, T. & Sigal, A. SARS-CoV-2 prolonged infection during advanced HIV disease evolves extensive immune escape. Cell Host & Microbe (2022). 10.1016/j.chom.2022.01.005

16 Karim, F., Moosa, M. Y., Gosnell, B., Sandile, C., Giandhari, J., Pillay, S., Tegally, H., Wilkinson, E., San, E. J. & Msomi, N. Persistent SARS-CoV-2 infection and intra-host evolution in association with advanced HIV infection. medRxiv (2021).

17 Riddell, A. C., Kele, B., Harris, K., Bible, J., Murphy, M., Dakshina, S., Storey, N., Owoyemi, D., Pade, C., Gibbons, J. M., Harrington, D., Alexander, E., McKnight, Á. & Cutino-Moguel, T. Generation of novel SARS-CoV-2 variants on B.1.1.7 lineage in three patients with advanced HIV disease. Clin Infect Dis (2022). PMC9213850. 10.1093/cid/ciac409

18 Kemp, S. A., Collier, D. A., Datir, R. P., Ferreira, I., Gayed, S., Jahun, A., Hosmillo, M., Rees-Spear, C., Mlcochova, P., Lumb, I. U., Roberts, D. J., Chandra, A., Temperton, N., Collaboration, C.-N. B. C.-., Consortium, C.-G. U., Sharrocks, K., Blane, E., Modis, Y., Leigh, K. E., Briggs, J. A. G., van Gils, M. J., Smith, K. G. C., Bradley, J. R., Smith, C., Doffinger, R., Ceron-Gutierrez, L., Barcenas-Morales, G., Pollock, D. D., Goldstein, R. A., Smielewska, A., Skittrall, J. P., Gouliouris, T., Goodfellow, I. G., Gkrania-Klotsas, E., Illingworth, C. J. R., McCoy, L. E. & Gupta, R. K. SARS-CoV-2 evolution during treatment of chronic infection. Nature 592, 277–282 (2021). PMC7610568. 10.1038/s41586-021-03291-y

19 Peacock, T. P., Penrice-Randal, R., Hiscox, J. A. & Barclay, W. S. SARS-CoV-2 one year on: evidence for ongoing viral adaptation. The Journal of General Virology 102 (2021).

20 Jensen, B., Luebke, N., Feldt, T., Keitel, V., Brandenburger, T., Kindgen-Milles, D., Lutterbeck, M., Freise, N., Schoeler, D. & Haas, R. Emergence of the E484K mutation in SARS-COV-2-infected immunocompromised patients treated with bamlanivimab in Germany. Lancet Reg Health Eur. 2021; 8: 100164. J. LANEPE (2021).

21 Baang, J. H., Smith, C., Mirabelli, C., Valesano, A. L., Manthei, D. M., Bachman, M. A., Wobus, C. E., Adams, M., Washer, L., Martin, E. T. & Lauring, A. S. Prolonged Severe Acute Respiratory Syndrome Coronavirus 2 Replication in an Immunocompromised Patient. J Infect Dis 223, 23–27 (2021). PMC7797758. 10.1093/infdis/jiaa666

22 Choi, B., Choudhary, M. C., Regan, J., Sparks, J. A., Padera, R. F., Qiu, X., Solomon, I. H., Kuo, H. H., Boucau, J., Bowman, K., Adhikari, U. D., Winkler, M. L., Mueller, A. A., Hsu, T. Y., Desjardins, M., Baden, L. R., Chan, B. T., Walker, B. D., Lichterfeld, M., Brigl, M., Kwon, D. S., Kanjilal, S., Richardson, E. T., Jonsson, A. H., Alter, G., Barczak, A. K., Hanage, W. P., Yu, X. G., Gaiha, G. D., Seaman, M. S., Cernadas, M. & Li, J. Z. Persistence and Evolution of SARS-CoV-2 in an Immunocompromised Host. N Engl J Med 383, 2291–2293 (2020). PMC7673303. 10.1056/NEJMc2031364

23 Avanzato, V. A., Matson, M. J., Seifert, S. N., Pryce, R., Williamson, B. N., Anzick, S. L., Barbian, K., Judson, S. D., Fischer, E. R., Martens, C., Bowden, T. A., de Wit, E., Riedo, F. X. & Munster, V. J. Case Study: Prolonged Infectious SARS-CoV-2 Shedding from an Asymptomatic Immunocompromised Individual with Cancer. Cell 183, 1901-1912.e1909 (2020). PMC7640888. 10.1016/j.cell.2020.10.049

24 Wilkinson, S. A. J., Richter, A., Casey, A., Osman, H., Mirza, J. D., Stockton, J., Quick, J., Ratcliffe, L., Sparks, N., Cumley, N., Poplawski, R., Nicholls, S. N., Kele, B., Harris, K., Peacock, T. P. & Loman, N. J. Recurrent SARS-CoV-2 mutations in immunodeficient patients. Virus Evol 8, veac050 (2022). PMC9384748. 10.1093/ve/veac050

25 Maponga, T. G., Jeffries, M., Tegally, H., Sutherland, A., Wilkinson, E., Lessells, R. J., Msomi, N., van Zyl, G., de Oliveira, T. & Preiser, W. Persistent SARS-CoV-2 infection with accumulation of mutations in a patient with poorly controlled HIV infection. Clin Infect Dis (2022). PMC9278209. 10.1093/cid/ciac548

26 Hoffman, S. A., Costales, C., Sahoo, M. K., Palanisamy, S., Yamamoto, F., Huang, C., Verghese, M., Solis, D. C., Sibai, M. & Subramanian, A. SARS-CoV-2 Neutralization Resistance Mutations in Patient with HIV/AIDS, California, USA. Emerging Infectious Diseases 27 (2021).

27 Corey, L., Beyrer, C., Cohen, M. S., Michael, N. L., Bedford, T. & Rolland, M. SARS-CoV-2 Variants in Patients with Immunosuppression. N Engl J Med 385, 562–566 (2021). PMC8494465. 10.1056/NEJMsb2104756

28 Kistler, K. E., Huddleston, J. & Bedford, T. Rapid and parallel adaptive mutations in spike S1 drive clade success in SARS-CoV-2. Cell Host Microbe 30, 545-555.e544 (2022). PMC8938189. 10.1016/j.chom.2022.03.018

29 Lauring, A. S., Jones, J. O. & Andino, R. Rationalizing the development of live attenuated virus vaccines. Nat Biotechnol 28, 573–579 (2010). PMC2883798. 10.1038/nbt.1635

30 Lamers, M. M. & Haagmans, B. L. SARS-CoV-2 pathogenesis. Nature Reviews Microbiology 20, 270–284 (2022). 10.1038/s41579-022-00713-0

31 Meng, B., Abdullahi, A., Ferreira, I. A. T. M., Goonawardane, N., Saito, A., Kimura, I., Yamasoba, D., Gerber, P. P., Fatihi, S., Rathore, S., Zepeda, S. K., Papa, G., Kemp, S. A., Ikeda, T., Toyoda, M., Tan, T. S., Kuramochi, J., Mitsunaga, S., Ueno, T., Shirakawa, K., Takaori-Kondo, A., Brevini, T., Mallery, D. L., Charles, O. J., Baker, S., Dougan, G., Hess, C., Kingston, N., Lehner, P. J., Lyons, P. A., Matheson, N. J., Owehand, W. H., Saunders, C., Summers, C., Thaventhiran, J. E. D., Toshner, M., Weekes, M. P., Maxwell, P., Shaw, A., Bucke, A., Calder, J., Canna, L., Domingo, J., Elmer, A., Fuller, S., Harris, J., Hewitt, S., Kennet, J., Jose, S., Kourampa, J., Meadows, A., O’Brien, C., Price, J., Publico, C., Rastall, R., Ribeiro, C., Rowlands, J., Ruffolo, V., Tordesillas, H., Bullman, B., Dunmore, B. J., Fawke, S., Gräf, S., Hodgson, J., Huang, C., Hunter, K., Jones, E., Legchenko, E., Matara, C., Martin, J., Mescia, F., O’Donnell, C., Pointon, L., Shih, J., Sutcliffe, R., Tilly, T., Treacy, C., Tong, Z., Wood, J., Wylot, M., Betancourt, A., Bower, G., Cossetti, C., De Sa, A., Epping, M., Fawke, S., Gleadall, N., Grenfell, R., Hinch, A., Jackson, S., Jarvis, I., Krishna, B., Nice, F., Omarjee, O., Perera, M., Potts, M., Richoz, N., Romashova, V., Stefanucci, L., Strezlecki, M., Turner, L., De Bie, E. M. D. D., Bunclark, K., Josipovic, M., Mackay, M., Allison, J., Butcher, H., Caputo, D., Clapham-Riley, D., Dewhurst, E., Furlong, A., Graves, B., Gray, J., Ivers, T., Le Gresley, E., Linger, R., Meloy, S., Muldoon, F., Ovington, N., Papadia, S., Phelan, I., Stark, H., Stirrups, K. E., Townsend, P., Walker, N., Webster, J., Scholtes, I., Hein, S., King, R., Butlertanaka, E. P., Tanaka, Y. L., Ikeda, T., Ito, J., Uriu, K., Kosugi, Y., Suganami, M., Oide, A., Yokoyama, M., Chiba, M., Motozono, C., Nasser, H., Shimizu, R., Yuan, Y., Kitazato, K., Hasebe, H., Nakagawa, S., Wu, J., Takahashi, M., Fukuhara, T., Shimizu, K., Tsushima, K., Kubo, H., Kazuma, Y., Nomura, R., Horisawa, Y., Nagata, K., Kawai, Y., Yanagida, Y., Tashiro, Y., Tokunaga, K., Ozono, S., Kawabata, R., Morizako, N., Sadamasu, K., Asakura, H., Nagashima, M., Yoshimura, K., Cárdenas, P., Muñoz, E., Barragan, V., Márquez, S., Prado-Vivar, B., Becerra-Wong, M., Caravajal, M., Trueba, G., Rojas-Silva, P., Grunauer, M., Gutierrez, B., Guadalupe, J. J., Fernández-Cadena, J. C., Andrade-Molina, D., Baldeon, M., Pinos, A., Bowen, J. E., Joshi, A., Walls, A. C., Jackson, L., Martin, D., Smith, K. G. C., Bradley, J., Briggs, J. A. G., Choi, J., Madissoon, E., Meyer, K., Mlcochova, P., Ceron-Gutierrez, L., Doffinger, R., Teichmann, S. A., Fisher, A. J., Pizzuto, M. S., de Marco, A., Corti, D., Hosmillo, M., Lee, J. H., James, L. C., Thukral, L., Veesler, D., Sigal, A., Sampaziotis, F., Goodfellow, I. G., Matheson, N. J., Sato, K., Gupta, R. K., The, C.-N. B. C.-C., The Genotype to Phenotype Japan Consortium, m. & Ecuador, C. C. Altered TMPRSS2 usage by SARS-CoV-2 Omicron impacts tropism and fusogenicity. Nature (2022). 10.1038/s41586-022-04474-x

32 Hoffmann, M., Kleine-Weber, H. & Pöhlmann, S. A Multibasic Cleavage Site in the Spike Protein of SARS-CoV-2 Is Essential for Infection of Human Lung Cells. Mol Cell 78, 779-784.e775 (2020). PMC7194065. 10.1016/j.molcel.2020.04.022

33 Papa, G., Mallery, D. L., Albecka, A., Welch, L. G., Cattin-Ortolá, J., Luptak, J., Paul, D., McMahon, H. T., Goodfellow, I. G., Carter, A., Munro, S. & James, L. C. Furin cleavage of SARS-CoV-2 Spike promotes but is not essential for infection and cell-cell fusion. PLoS Pathog 17, e1009246 (2021). PMC7861537. 10.1371/journal.ppat.1009246

34 Cattin-Ortolá, J., Welch, L. G., Maslen, S. L., Papa, G., James, L. C. & Munro, S. Sequences in the cytoplasmic tail of SARS-CoV-2 Spike facilitate expression at the cell surface and syncytia formation. Nat Commun 12, 5333 (2021). PMC8429659. 10.1038/s41467-021-25589-1

35 Buchrieser, J., Dufloo, J., Hubert, M., Monel, B., Planas, D., Rajah, M. M., Planchais, C., Porrot, F., Guivel-Benhassine, F., Van der Werf, S., Casartelli, N., Mouquet, H., Bruel, T. & Schwartz, O. Syncytia formation by SARS-CoV-2-infected cells. Embo j 39, e106267 (2020). PMC7646020. 10.15252/embj.2020106267

36 Braga, L., Ali, H., Secco, I., Chiavacci, E., Neves, G., Goldhill, D., Penn, R., Jimenez-Guardeño, J. M., Ortega-Prieto, A. M., Bussani, R., Cannatà, A., Rizzari, G., Collesi, C., Schneider, E., Arosio, D., Shah, A. M., Barclay, W. S., Malim, M. H., Burrone, J. & Giacca, M. Drugs that inhibit TMEM16 proteins block SARS-CoV-2 spike-induced syncytia. Nature 594, 88–93 (2021). PMC7611055. 10.1038/s41586-021-03491-6

37 Zhang, Z., Zheng, Y., Niu, Z., Zhang, B., Wang, C., Yao, X., Peng, H., Franca, D. N., Wang, Y., Zhu, Y., Su, Y., Tang, M., Jiang, X., Ren, H., He, M., Wang, Y., Gao, L., Zhao, P., Shi, H., Chen, Z., Wang, X., Piacentini, M., Bian, X., Melino, G., Liu, L., Huang, H. & Sun, Q. SARS-CoV-2 spike protein dictates syncytium-mediated lymphocyte elimination. Cell Death Differ 28, 2765–2777 (2021). PMC8056997. 10.1038/s41418-021-00782-3

38 Bussani, R., Schneider, E., Zentilin, L., Collesi, C., Ali, H., Braga, L., Volpe, M. C., Colliva, A., Zanconati, F., Berlot, G., Silvestri, F., Zacchigna, S. & Giacca, M. Persistence of viral RNA, pneumocyte syncytia and thrombosis are hallmarks of advanced COVID-19 pathology. EBioMedicine 61, 103104 (2020). PMC7677597. 10.1016/j.ebiom.2020.103104

39 Hoffmann, M., Kleine-Weber, H., Schroeder, S., Krüger, N., Herrler, T., Erichsen, S., Schiergens, T. S., Herrler, G., Wu, N. H., Nitsche, A., Müller, M. A., Drosten, C. & Pöhlmann, S. SARS-CoV-2 Cell Entry Depends on ACE2 and TMPRSS2 and Is Blocked by a Clinically Proven Protease Inhibitor. Cell 181, 271-280.e278 (2020). PMC7102627. 10.1016/j.cell.2020.02.052

40 Peacock, T. P., Brown, J. C., Zhou, J., Thakur, N., Sukhova, K., Newman, J., Kugathasan, R., Yan, A. W. C., Furnon, W., De Lorenzo, G., Cowton, V. M., Reuss, D., Moshe, M., Quantrill, J. L., Platt, O. K., Kaforou, M., Patel, A. H., Palmarini, M., Bailey, D. & Barclay, W. S. The altered entry pathway and antigenic distance of the SARS-CoV-2 Omicron variant map to separate domains of spike protein. bioRxiv, 2021.2012.2031.474653 (2022). 10.1101/2021.12.31.474653

41 Willett, B. J., Grove, J., MacLean, O. A., Wilkie, C., De Lorenzo, G., Furnon, W., Cantoni, D., Scott, S., Logan, N., Ashraf, S., Manali, M., Szemiel, A., Cowton, V., Vink, E., Harvey, W. T., Davis, C., Asamaphan, P., Smollett, K., Tong, L., Orton, R., Hughes, J., Holland, P., Silva, V., Pascall, D. J., Puxty, K., da Silva Filipe, A., Yebra, G., Shaaban, S., Holden, M. T. G., Pinto, R. M., Gunson, R., Templeton, K., Murcia, P. R., Patel, A. H., Klenerman, P., Dunachie, S., Dunachie, S., Klenerman, P., Barnes, E., Brown, A., Adele, S., Kronsteiner, B., Murray, S. M., Abraham, P., Deeks, A., Ansari, M. A., de Silva, T., Turtle, L., Moore, S., Austin, J., Richter, A., Duncan, C., Payne, R., Ash, A., Koshy, C., Kele, B., Cutino-Moguel, T., Fairley, D. J., McKenna, J. P., Curran, T., Adams, H., Fraser, C., Bonsall, D., Fryer, H., Lythgoe, K., Thomson, L., Golubchik, T., Murray, A., Singleton, D., Beckwith, S. M., Mantzouratou, A., Barrow, M., Buchan, S. L., Reynolds, N., Warne, B., Maksimovic, J., Spellman, K., McCluggage, K., John, M., Beer, R., Afifi, S., Morgan, S., Mack, A., Marchbank, A., Price, A., Morriss, A., Bresner, C., Kitchen, C., Merrick, I., Southgate, J., Guest, M., Jones, O., Munn, R., Connor, T. R., Whalley, T., Workman, T., Fuller, W., Patel, A., Patel, B., Nebbia, G., Edgeworth, J., Snell, L. B., Batra, R., Charalampous, T., Beckett, A. H., Shelest, E., Robson, S. C., Underwood, A. P., Taylor, B. E. W., Yeats, C. A., Aanensen, D. M., Abudahab, K., Menegazzo, M., Joseph, A., Clark, G., Howson-Wells, H. C., Berry, L., Khakh, M., Lister, M. M., Boswell, T., Fleming, V. M., Holmes, C. W., McMurray, C. L., Shaw, J., Tang, J. W., Fallon, K., Odedra, M., Willford, N. J., Bird, P. W., Helmer, T., Williams, L.-A., Sheriff, N., Campbell, S., Raviprakash, V., Blakey, V., Moore, C., Sang, F., Debebe, J., Carlile, M., Loose, M. W., Holmes, N., Wright, V., Torok, M. E., Hamilton, W. L., Carabelli, A. M., Jermy, A., Blane, B., Churcher, C. M., Ludden, C., Aggarwal, D., Westwick, E., Brooks, E., McManus, G. M., Galai, K., Smith, K., Smith, K. S., Cox, M., Fragakis, M., Maxwell, P., Judges, S., Peacock, S. J., Feltwell, T., Kenyon, A., Eldirdiri, S., Davis, T., Taylor, J. F., Tan, N. K., Zarebski, A. E., Gutierrez, B., Raghwani, J., du Plessis, L., Kraemer, M. U. G., Pybus, O. G., Francois, S., Attwood, S. W., Vasylyeva, T. I., Jahun, A. S., Goodfellow, I. G., Georgana, I., Pinckert, M. L., Hosmillo, M., Izuagbe, R., Chaudhry, Y., Ryan, F., Lowe, H., Moses, S., Bedford, L., Cargill, J. S., Hughes, W., Moore, J., Stonehouse, S., Shah, D., Lee, J. C. D., Brown, J. R., Harris, K. A., Atkinson, L., Storey, N., Spyer, M. J., Flaviani, F., Alcolea-Medina, A., Sehmi, J., Ramble, J., Ohemeng-Kumi, N., Smith, P., Bertolusso, B., Thomas, C., Vernet, G., Lynch, J., Moore, N., Cortes, N., Williams, R., Kidd, S. P., Levett, L. J., Pusok, M., Grant, P. R., Kirk, S., Chatterton, W., Xu-McCrae, L., Smith, D. L., Young, G. R., Bashton, M., Kitchman, K., Gajee, K., Eastick, K., Lillie, P. J., Burns, P. J., Everson, W., Cox, A., Holmes, A. H., Bolt, F., Price, J. R., Pond, M., Randell, P. A., Madona, P., Mookerjee, S., Volz, E. M., Geidelberg, L., Ragonnet-Cronin, M., Boyd, O., Johnson, R., Pope, C. F., Witney, A. A., Monahan, I. M., Laing, K. G., Smollett, K. L., McNally, A., McMurray, C., Stockton, J., Quick, J., Loman, N. J., Poplawski, R., Nicholls, S., Rowe, W., Castigador, A., Macnaughton, E., Bouzidi, K. E., Sudhanva, M., Lampejo, T., Martinez Nunez, R. T., Breen, C., Sluga, G., Withell, K. T., Machin, N. W., George, R. P., Ahmad, S. S. Y., Pritchard, D. T., Binns, D., Wong, N., James, V., Williams, C., Illingworth, C. J., Jackson, C., de Angelis, D., Pascall, D., Mukaddas, A., Broos, A., da Silva Filipe, A., Mair, D., Robertson, D. L., Wright, D. W., Thomson, E. C., Starinskij, I., Tsatsani, I., Shepherd, J. G., Nichols, J., Hughes, J., Nomikou, K., Tong, L., Orton, R. J., Vattipally, S., Harvey, W. T., Sanderson, R., O’Brien, S., Rushton, S., Perkins, J., Blacow, R., Gunson, R. N., Gallagher, A., Consortium, P. & The, C.-G. U. K. C. SARS-CoV-2 Omicron is an immune escape variant with an altered cell entry pathway. Nature Microbiology 7, 1161–1179 (2022). 10.1038/s41564-022-01143-7

42 Saito, A., Irie, T., Suzuki, R., Maemura, T., Nasser, H., Uriu, K., Kosugi, Y., Shirakawa, K., Sadamasu, K., Kimura, I., Ito, J., Wu, J., Iwatsuki-Horimoto, K., Ito, M., Yamayoshi, S., Loeber, S., Tsuda, M., Wang, L., Ozono, S., Butlertanaka, E. P., Tanaka, Y. L., Shimizu, R., Shimizu, K., Yoshimatsu, K., Kawabata, R., Sakaguchi, T., Tokunaga, K., Yoshida, I., Asakura, H., Nagashima, M., Kazuma, Y., Nomura, R., Horisawa, Y., Yoshimura, K., Takaori-Kondo, A., Imai, M., Tanaka, S., Nakagawa, S., Ikeda, T., Fukuhara, T., Kawaoka, Y. & Sato, K. Enhanced fusogenicity and pathogenicity of SARS-CoV-2 Delta P681R mutation. Nature 602, 300–306 (2022). PMC8828475. 10.1038/s41586-021-04266-9

43 Rajah, M. M., Hubert, M., Bishop, E., Saunders, N., Robinot, R., Grzelak, L., Planas, D., Dufloo, J., Gellenoncourt, S., Bongers, A., Zivaljic, M., Planchais, C., Guivel-Benhassine, F., Porrot, F., Mouquet, H., Chakrabarti, L. A., Buchrieser, J. & Schwartz, O. SARS-CoV-2 Alpha, Beta, and Delta variants display enhanced Spike-mediated syncytia formation. Embo j 40, e108944 (2021). PMC8646911. 10.15252/embj.2021108944

44 Bowen, J. E., Addetia, A., Dang, H. V., Stewart, C., Brown, J. T., Sharkey, W. K., Sprouse, K. R., Walls, A. C., Mazzitelli, I. G., Logue, J. K., Franko, N. M., Czudnochowski, N., Powell, A. E., Dellota, E., Jr., Ahmed, K., Ansari, A. S., Cameroni, E., Gori, A., Bandera, A., Posavad, C. M., Dan, J. M., Zhang, Z., Weiskopf, D., Sette, A., Crotty, S., Iqbal, N. T., Corti, D., Geffner, J., Snell, G., Grifantini, R., Chu, H. Y. & Veesler, D. Omicron spike function and neutralizing activity elicited by a comprehensive panel of vaccines. Science 377, 890–894 (2022). PMC9348749. 10.1126/science.abq0203

45 Mlcochova, P., Kemp, S. A., Dhar, M. S., Papa, G., Meng, B., Ferreira, I., Datir, R., Collier, D. A., Albecka, A., Singh, S., Pandey, R., Brown, J., Zhou, J., Goonawardane, N., Mishra, S., Whittaker, C., Mellan, T., Marwal, R., Datta, M., Sengupta, S., Ponnusamy, K., Radhakrishnan, V. S., Abdullahi, A., Charles, O., Chattopadhyay, P., Devi, P., Caputo, D., Peacock, T., Wattal, C., Goel, N., Satwik, A., Vaishya, R., Agarwal, M., Mavousian, A., Lee, J. H., Bassi, J., Silacci-Fegni, C., Saliba, C., Pinto, D., Irie, T., Yoshida, I., Hamilton, W. L., Sato, K., Bhatt, S., Flaxman, S., James, L. C., Corti, D., Piccoli, L., Barclay, W. S., Rakshit, P., Agrawal, A. & Gupta, R. K. SARS-CoV-2 B.1.617.2 Delta variant replication and immune evasion. Nature 599, 114–119 (2021). PMC8566220. 10.1038/s41586-021-03944-y

46 Kimura, I., Yamasoba, D., Tamura, T., Nao, N., Suzuki, T., Oda, Y., Mitoma, S., Ito, J., Nasser, H., Zahradnik, J., Uriu, K., Fujita, S., Kosugi, Y., Wang, L., Tsuda, M., Kishimoto, M., Ito, H., Suzuki, R., Shimizu, R., Begum, M. M., Yoshimatsu, K., Kimura, K. T., Sasaki, J., Sasaki-Tabata, K., Yamamoto, Y., Nagamoto, T., Kanamune, J., Kobiyama, K., Asakura, H., Nagashima, M., Sadamasu, K., Yoshimura, K., Shirakawa, K., Takaori-Kondo, A., Kuramochi, J., Schreiber, G., Ishii, K. J., Hashiguchi, T., Ikeda, T., Saito, A., Fukuhara, T., Tanaka, S., Matsuno, K. & Sato, K. Virological characteristics of the SARS-CoV-2 Omicron BA.2 subvariants, including BA.4 and BA.5. Cell (2022). PMC9472642. 10.1016/j.cell.2022.09.018

47 Willett, B. J., Grove, J., MacLean, O. A., Wilkie, C., De Lorenzo, G., Furnon, W., Cantoni, D., Scott, S., Logan, N., Ashraf, S., Manali, M., Szemiel, A., Cowton, V., Vink, E., Harvey, W. T., Davis, C., Asamaphan, P., Smollett, K., Tong, L., Orton, R., Hughes, J., Holland, P., Silva, V., Pascall, D. J., Puxty, K., da Silva Filipe, A., Yebra, G., Shaaban, S., Holden, M. T. G., Pinto, R. M., Gunson, R., Templeton, K., Murcia, P. R., Patel, A. H., Klenerman, P., Dunachie, S., Haughney, J., Robertson, D. L., Palmarini, M., Ray, S. & Thomson, E. C. SARS-CoV-2 Omicron is an immune escape variant with an altered cell entry pathway. Nat Microbiol 7, 1161–1179 (2022). PMC9352574 COVID-19. The other authors declare no competing interests. 10.1038/s41564-022-01143-7

48 Katsura, H., Sontake, V., Tata, A., Kobayashi, Y., Edwards, C. E., Heaton, B. E., Konkimalla, A., Asakura, T., Mikami, Y., Fritch, E. J., Lee, P. J., Heaton, N. S., Boucher, R. C., Randell, S. H., Baric, R. S. & Tata, P. R. Human Lung Stem Cell-Based Alveolospheres Provide Insights into SARS-CoV-2-Mediated Interferon Responses and Pneumocyte Dysfunction. Cell Stem Cell 27, 890-904.e898 (2020). PMC7577733. 10.1016/j.stem.2020.10.005

49 Delorey, T. M., Ziegler, C. G. K., Heimberg, G., Normand, R., Yang, Y., Segerstolpe, Å., Abbondanza, D., Fleming, S. J., Subramanian, A., Montoro, D. T., Jagadeesh, K. A., Dey, K. K., Sen, P., Slyper, M., Pita-Juárez, Y. H., Phillips, D., Biermann, J., Bloom-Ackermann, Z., Barkas, N., Ganna, A., Gomez, J., Melms, J. C., Katsyv, I., Normandin, E., Naderi, P., Popov, Y. V., Raju, S. S., Niezen, S., Tsai, L. T., Siddle, K. J., Sud, M., Tran, V. M., Vellarikkal, S. K., Wang, Y., Amir-Zilberstein, L., Atri, D. S., Beechem, J., Brook, O. R., Chen, J., Divakar, P., Dorceus, P., Engreitz, J. M., Essene, A., Fitzgerald, D. M., Fropf, R., Gazal, S., Gould, J., Grzyb, J., Harvey, T., Hecht, J., Hether, T., Jané-Valbuena, J., Leney-Greene, M., Ma, H., McCabe, C., McLoughlin, D. E., Miller, E. M., Muus, C., Niemi, M., Padera, R., Pan, L., Pant, D., Pe’er, C., Pfiffner-Borges, J., Pinto, C. J., Plaisted, J., Reeves, J., Ross, M., Rudy, M., Rueckert, E. H., Siciliano, M., Sturm, A., Todres, E., Waghray, A., Warren, S., Zhang, S., Zollinger, D. R., Cosimi, L., Gupta, R. M., Hacohen, N., Hibshoosh, H., Hide, W., Price, A. L., Rajagopal, J., Tata, P. R., Riedel, S., Szabo, G., Tickle, T. L., Ellinor, P. T., Hung, D., Sabeti, P. C., Novak, R., Rogers, R., Ingber, D. E., Jiang, Z. G., Juric, D., Babadi, M., Farhi, S. L., Izar, B., Stone, J. R., Vlachos, I. S., Solomon, I. H., Ashenberg, O., Porter, C. B. M., Li, B., Shalek, A. K., Villani, A. C., Rozenblatt-Rosen, O. & Regev, A. COVID-19 tissue atlases reveal SARS-CoV-2 pathology and cellular targets. Nature 595, 107–113 (2021). PMC8919505. 10.1038/s41586-021-03570-8

50 Zhu, N., Wang, W., Liu, Z., Liang, C., Wang, W., Ye, F., Huang, B., Zhao, L., Wang, H., Zhou, W., Deng, Y., Mao, L., Su, C., Qiang, G., Jiang, T., Zhao, J., Wu, G., Song, J. & Tan, W. Morphogenesis and cytopathic effect of SARS-CoV-2 infection in human airway epithelial cells. Nat Commun 11, 3910 (2020). PMC7413383. 10.1038/s41467-020-17796-z

51 Li, S., Zhang, Y., Guan, Z., Li, H., Ye, M., Chen, X., Shen, J., Zhou, Y., Shi, Z. L., Zhou, P. & Peng, K. SARS-CoV-2 triggers inflammatory responses and cell death through caspase-8 activation. Signal Transduct Target Ther 5, 235 (2020). PMC7545816. 10.1038/s41392-020-00334-0

52 Ren, Y., Shu, T., Wu, D., Mu, J., Wang, C., Huang, M., Han, Y., Zhang, X. Y., Zhou, W., Qiu, Y. & Zhou, X. The ORF3a protein of SARS-CoV-2 induces apoptosis in cells. Cell Mol Immunol 17, 881–883 (2020). PMC7301057. 10.1038/s41423-020-0485-9

53 Chan, F. K., Luz, N. F. & Moriwaki, K. Programmed necrosis in the cross talk of cell death and inflammation. Annu Rev Immunol 33, 79–106 (2015). PMC4394030. 10.1146/annurev-immunol-032414-112248

54 Rock, K. L. & Kono, H. The inflammatory response to cell death. Annu Rev Pathol 3, 99–126 (2008). PMC3094097. 10.1146/annurev.pathmechdis.3.121806.151456

55 Zhu, Z., Shi, J., Li, L., Wang, J., Zhao, Y. & Ma, H. Therapy Targets SARS-CoV-2 Infection-Induced Cell Death. Front Immunol 13, 870216 (2022). PMC9152132. 10.3389/fimmu.2022.870216

56 Karim, F., Gazy, I., Cele, S., Zungu, Y., Krause, R., Bernstein, M., Khan, K., Ganga, Y., Rodel, H. E., Mthabela, N., Mazibuko, M., Muema, D., Ramjit, D., Ndung’u, T., Hanekom, W., Gosnell, B., Team, C.-K., Lessells, R. J., Wong, E. B., de Oliveira, T., Moosa, Y., Lustig, G., Leslie, A., Kloverpris, H. & Sigal, A. HIV status alters disease severity and immune cell responses in beta variant SARS-CoV-2 infection wave. Elife 10 (2021). 10.7554/eLife.67397

57 Greaney, A. J., Loes, A. N., Crawford, K. H. D., Starr, T. N., Malone, K. D., Chu, H. Y. & Bloom, J. D. Comprehensive mapping of mutations in the SARS-CoV-2 receptor-binding domain that affect recognition by polyclonal human plasma antibodies. Cell Host Microbe 29, 463-476.e466 (2021). PMC7869748. 10.1016/j.chom.2021.02.003

58 Cele, S., Jackson, L., Khoury, D. S., Khan, K., Moyo-Gwete, T., Tegally, H., San, J. E., Cromer, D., Scheepers, C., Amoako, D. G., Karim, F., Bernstein, M., Lustig, G., Archary, D., Smith, M., Ganga, Y., Jule, Z., Reedoy, K., Hwa, S. H., Giandhari, J., Blackburn, J. M., Gosnell, B. I., Abdool Karim, S. S., Hanekom, W., von Gottberg, A., Bhiman, J. N., Lessells, R. J., Moosa, M. S., Davenport, M. P., de Oliveira, T., Moore, P. L. & Sigal, A. Omicron extensively but incompletely escapes Pfizer BNT162b2 neutralization. Nature (2021). 10.1038/s41586-021-04387-1

59 Sigal, A., Danon, T., Cohen, A., Milo, R., Geva-Zatorsky, N., Lustig, G., Liron, Y., Alon, U. & Perzov, N. Generation of a fluorescently labeled endogenous protein library in living human cells. Nat Protoc 2, 1515–1527 (2007). 10.1038/nprot.2007.197

60 Jackson, L., Hunter, J., Cele, S., Ferreira, I. M., Young, A. C., Karim, F., Madansein, R., Dullabh, K. J., Chen, C. Y., Buckels, N. J., Ganga, Y., Khan, K., Boulle, M., Lustig, G., Neher, R. A. & Sigal, A. Incomplete inhibition of HIV infection results in more HIV infected lymph node cells by reducing cell death. Elife 7 (2018). PMC5896883. 10.7554/eLife.30134

61 Sigal, A., Kim, J. T., Balazs, A. B., Dekel, E., Mayo, A., Milo, R. & Baltimore, D. Cell-to-cell spread of HIV permits ongoing replication despite antiretroviral therapy. Nature 477, 95–98 (2011). 10.1038/nature10347

62 Ko, S. H., Bayat Mokhtari, E., Mudvari, P., Stein, S., Stringham, C. D., Wagner, D., Ramelli, S., Ramos-Benitez, M. J., Strich, J. R., Davey, R. T., Jr., Zhou, T., Misasi, J., Kwong, P. D., Chertow, D. S., Sullivan, N. J. & Boritz, E. A. High-throughput, single-copy sequencing reveals SARS-CoV-2 spike variants coincident with mounting humoral immunity during acute COVID-19. PLoS Pathog 17, e1009431 (2021). PMC8031304. 10.1371/journal.ppat.1009431

63 Jackson, L., Rodel, H., Hwa, S.-H., Cele, S., Ganga, Y., Tegally, H., Bernstein, M., Giandhari, J., Gosnell, B. I., Khan, K., Hanekom, W., Karim, F., de Oliveira, T., Moosa, M.-Y. S. & Sigal, A. SARS-CoV-2 cell-to-cell spread occurs rapidly and is insensitive to antibody neutralization. bioRxiv, 2021.2006.2001.446516 (2021). 10.1101/2021.06.01.446516

64 Rajah, M. M., Bernier, A., Buchrieser, J. & Schwartz, O. The Mechanism and Consequences of SARS-CoV-2 Spike-Mediated Fusion and Syncytia Formation. J Mol Biol 434, 167280 (2022). PMC8485708. 10.1016/j.jmb.2021.167280

65 Mlcochova, P., Kemp, S., Dhar, M. S., Papa, G., Meng, B., Ferreira, I., Datir, R., Collier, D. A., Albecka, A., Singh, S., Pandey, R., Brown, J., Zhou, J., Goonawardane, N., Mishra, S., Whittaker, C., Mellan, T., Marwal, R., Datta, M., Sengupta, S., Ponnusamy, K., Radhakrishnan, V. S., Abdullahi, A., Charles, O., Chattopadhyay, P., Devi, P., Caputo, D., Peacock, T., Wattal, D. C., Goel, N., Satwik, A., Vaishya, R., Agarwal, M., Indian, S.-C.-G. C., Genotype to Phenotype Japan, C., Collaboration, C.-N. B. C.-., Mavousian, A., Lee, J. H., Bassi, J., Silacci-Fegni, C., Saliba, C., Pinto, D., Irie, T., Yoshida, I., Hamilton, W. L., Sato, K., Bhatt, S., Flaxman, S., James, L. C., Corti, D., Piccoli, L., Barclay, W. S., Rakshit, P., Agrawal, A. & Gupta, R. K. SARS-CoV-2 B.1.617.2 Delta variant replication and immune evasion. Nature (2021). 10.1038/s41586-021-03944-y

66 Nardacci, R., Perfettini, J. L., Grieco, L., Thieffry, D., Kroemer, G. & Piacentini, M. Syncytial apoptosis signaling network induced by the HIV-1 envelope glycoprotein complex: an overview. Cell Death Dis 6, e1846 (2015). PMC4558497. 10.1038/cddis.2015.204

67 Wolter, N., Jassat, W., Walaza, S., Welch, R., Moultrie, H., Groome, M., Amoako, D. G., Everatt, J., Bhiman, J. N., Scheepers, C., Tebeila, N., Chiwandire, N., du Plessis, M., Govender, N., Ismail, A., Glass, A., Mlisana, K., Stevens, W., Treurnicht, F. K., Makatini, Z., Hsiao, N. Y., Parboosing, R., Wadula, J., Hussey, H., Davies, M. A., Boulle, A., von Gottberg, A. & Cohen, C. Early assessment of the clinical severity of the SARS-CoV-2 omicron variant in South Africa: a data linkage study. Lancet 399, 437–446 (2022). PMC8769664 Medical Research Council, the UK Foreign, Commonwealth and Development Office, the Wellcome Trust, the US Centers for Disease Control and Prevention, and Sanofi Pasteur. NW and MdP have received grant support from Sanofi Pasteur and the Bill & Melinda Gates Foundation. AvG has received grant support from Sanofi Pasteur, the US Centers for Disease Control and Prevention, the South African Medical Research Council, the Bill & Melinda Gates Foundation, WHO, the Fleming Fund, and the Wellcome Trust. RW declares personal shareholding in Adcock Ingram Holdings, Dischem Pharmacies, Discovery, Netcare, and Aspen Pharmacare Holdings. WS has received grant support from the South African Medical Research Council, with funds received from the Department of Science and Innovation, and the Bill & Melinda Gates Foundation. All other authors declare no competing interests. 10.1016/s0140-6736(22)00017-4

68 Khan, K., Karim, F., Cele, S., Reedoy, K., San, J. E., Lustig, G., Tegally, H., Rosenberg, Y., Bernstein, M., Jule, Z., Ganga, Y., Ngcobo, N., Mazibuko, M., Mthabela, N., Mhlane, Z., Mbatha, N., Miya, Y., Giandhari, J., Ramphal, Y., Naidoo, T., Sivro, A., Samsunder, N., Kharsany, A. B. M., Amoako, D., Bhiman, J. N., Manickchund, N., Karim, Q. A., Magula, N., Abdool Karim, S. S., Gray, G., Hanekom, W., von Gottberg, A., Milo, R., Gosnell, B. I., Lessells, R. J., Moore, P. L., de Olveira, T., Moosa, M. S. & Sigal, A. Omicron infection enhances Delta antibody immunity in vaccinated persons. Nature (2022). 10.1038/s41586-022-04830-x

69 Cele, S., Gazy, I., Jackson, L., Hwa, S. H., Tegally, H., Lustig, G., Giandhari, J., Pillay, S., Wilkinson, E., Naidoo, Y., Karim, F., Ganga, Y., Khan, K., Bernstein, M., Balazs, A. B., Gosnell, B. I., Hanekom, W., Moosa, M. S., Network for Genomic Surveillance in South, A., Team, C.-K., Lessells, R. J., de Oliveira, T. & Sigal, A. Escape of SARS-CoV-2 501Y.V2 from neutralization by convalescent plasma. Nature 593, 142–146 (2021). 10.1038/s41586-021-03471-w

